# Performance of an artificial intelligence foundation model for prostate radiotherapy segmentation

**DOI:** 10.1101/2025.02.23.25322754

**Authors:** Matt Doucette, Chien-Yi Liao, Mu-Han Lin, Steve Jiang, Dan Nguyen, Daniel X. Yang

## Abstract

**Importance:** Artificial intelligence (AI) foundation models such as Segment Anything Model 2 (SAM 2) offer potential for semi-automated image segmentation with minimal fine-tuning, but their performance in specialized clinical tasks like radiation therapy planning are not well characterized.

**Objective:** To evaluate the performance of SAM 2 in segmenting pre-operative intact prostate and post-operative prostate fossa targets for prostate radiotherapy planning.

**Design, Setting, Participants:** Retrospective cohort study deploying and testing a foundation model for AI segmentation for prostate radiotherapy planning. CT simulation images and radiation plans were obtained from a single academic institution for patients undergoing prostate cancer treatment. Data analysis was performed from September 2024 to February 2025.

**Exposures:** AI segmentation with varying levels of human intervention, ranging from intervals of every 2nd to every 10th ground truth slice provided as input.

**Main Outcome and Measures:** Segmentation accuracy measured by Dice Similarity Coefficient (DSC) and Hausdorff Distance (HD) for intact and post-operative prostate target delineation.

**Results:** While SAM 2 outperformed interpolation in DSC and HD for both intact and post-operative prostate cancer patient cases, the AI segmentation accuracy was significantly better in the intact pre-operative patient cases where anatomic boundaries were better defined than post-operative patient cases. This is especially evident when sparse ground truth was provided simulating lower levels of human intervention.

**Conclusions and Relevance:** AI foundation models show promising application for specialized medical tasks such as prostate cancer radiotherapy segmentation with limited need for fine-tuning or retraining, although their clinical application will require further understanding of task-specific performance.

## Introduction

In recent years, the development of robust machine learning models has been a resource-intensive endeavor, requiring significant computational power, large-scale annotated datasets, and substantial funding.^1–3^ The emergence of foundation models has introduced new toolsets to this landscape, enabling researchers and practitioners to leverage pre-trained models and fine-tune them for specialized applications.^4–8^ These foundation models, such as Meta’s Segment Anything Model 2 (SAM 2) and OpenAI’s ChatGPT, have demonstrated versatility across diverse domains, including medical imaging and natural language processing. Applying foundation models to specialized clinical and research tasks may offset the prohibitive costs associated with training such large-scale models from scratch.^9–12^

Radiation therapy is a major treatment modality for localized prostate cancer. Accurate segmentation for radiation treatment planning is crucial, as it directly impacts the precision of the treatment delivered and patient outcomes.^13,14^ While artificial intelligence (AI) tools have been introduced to aid segmentation, it remains a labor-intensive task requiring expert clinician input. AI auto-segmentation models can also be challenging to implement clinically due to data heterogeneity, lack of standardization across practice settings, variations in patient anatomy and tumor boundaries, hurdles in clinical workflow integration, amongst other factors.^15,16^ Foundation models have been postulated to help address these challenges, potentially reducing time and costs without extensive model retraining efforts.^17,18^

In this study, we aim to study an open-source foundation model designed to segment objects in various contexts with minimal user input. While initially developed for general-purpose segmentation tasks, the model’s adaptability has sparked interest in its application to medical imaging, including radiotherapy planning.^19,20^ Specifically, we aim to characterize the segmentation performance of SAM 2 in pre-operative intact prostate cancer and post-operative prostate fossa cancer cases. We subsequently assess SAM 2’s capability to perform effectively with varying levels of human intervention by providing sparse ground truth annotations.

By applying a foundation model to radiotherapy segmentation, this work highlights the potential for future foundation models to assist clinicians in specialized medical segmentation tasks, particularly in scenarios with limited resources or minimal expert input. The findings could have broader implications for the adoption of foundation models in medical imaging and clinical practice.

## Methods

### RT Planning and Data Acquisition

This study included 282 prostate cancer patient cases from a single academic institution, divided into pre-operative (N=139) and post-operative (N=143) cohorts. The pre-operative patient cases include the clinical target volume (CTV) of the intact prostate, while the post-operative cases include the CTV of the prostate fossa following prostatectomy. Data was retrospectively collected and includes CT volumes obtained during simulation and treatment planning for definitive radiotherapy to the prostate and for adjuvant or salvage post-operative radiotherapy to the prostatic fossa.^21^ All patients underwent CT imaging for simulation and treatment planning between 2021 and 2024.^22^ The CTVs range from 10 to 39 slices across all the pre-operative and post-operative cases. The voxel size of the CT images is 1.17 × 1.17 × 2 mm^3^, ensuring sufficient resolution for delineating treatment targets and surrounding structures.

### Model and Input

Raw DICOM data was parsed to create a folder with the JPG images and corresponding three-dimensional binary mask of the anatomical structure of interest (intact prostate or prostate fossa). The raw DICOM data was normalized using a sigmoid function transformation with α = 0.02 to create images that maximize the dynamic range near the average pixel value of the structure of interest.^23^ There are 4 different-sized SAM 2 models called tiny, small, base plus, and large with sizes of 38.9, 46, 80.8, and 224.4 Mb respectively. The large model was used, which offers the best performance at the slowest inference rate.

### Experiment Design

The SAM 2 model supports multiple input modalities, including positive and negative clicks, bounding boxes, and masks.^9^ Input masks were used for this study. For each case, the physician-created CTV contour was used as the ground truth segmentation. A subset of these ground truth slices was provided as input to the SAM 2 model, simulating how a human clinician would typically segment every few slices in clinical practice and subsequently interpolate to create the CTV. In this study, the first and last ground truth slices were always included, with additional ground truth slices supplied at intervals to emulate different levels of human intervention.

For example, in a case where the CTV contains 14 slices total, using an interval of every 4th slice would mean the 1st, 4th, 8th, 12th, and 14th slices were provided as input to the SAM 2 model. The remaining slices (2nd, 3rd, 5th, 6th, 7th, 9th, 10th, 11th, and 13th) were predicted by the model. CTV segmentation accuracy for intervals ranging from every 2nd to every 10th slice were evaluated for each patient case using the Dice Similarity Coefficient (DSC) and Hausdorff

Distance (HD). To serve as a baseline comparison to SAM 2 performance, the missing slices were estimated using the interpolation of the given ground truth slices. The interpolated slices were calculated as the weighted linear combination of the Euclidean distance transforms of the last given and next given ground truth masks.^24^

The average SAM 2 segmentation DSC scores across superior-inferior anatomical positions for both pre-operative (prostate) and post-operative (prostate fossa) cases were graphed and visualized to better understand how anatomic location influenced model performance.

### Statistical Analysis

Performance comparisons were conducted at each level of ground truth provided (different levels of human intervention). Within each cohort, the performance of SAM 2 was compared to that of interpolation for intervals ranging from every 2nd to every 10th slice given. To assess statistically significant differences in the DSC and HD scores, the Mann-Whitney U test was used.^25^ Statistical analysis was performed using the scipy.stats module in Python 3.11.3.

## Results

### Patient and Treatment Characteristics

The pre-operative patient cohort had a median age of 65 years (IQR 56-74) and the post-operative cohort had a median age of 64 (IQR 54-76). Across both clinical settings, patients underwent simulation and treatment between 2021 to 2024. CTV volume was on average 40.22 cm^3^ (STD: 16.89 cm^3^) in the pre-operative setting compared to 146.51 cm^3^ (STD: 45.78 cm^3^) in the post-operative setting (Table 1).

**Table 1.** Patient Characteristics.

| Variable | Pre-op (N=139) | Post-op (N=143) |
| --- | --- | --- |
| Median Age (Range) | 65 (45–80) | 64 (40–88) |
| Year of simulation | 20021–2022 | 2022–2024 |
| CTV volume in cm<sup>3</sup> (mean ± SD) | 40.22 ± 16.89 | 146.51 ± 45.78 |

### Performance Trends

As the interval between provided ground truth slices increased, the performance of the SAM 2 model declined in both pre-operative and post-operative patient cases (Figure 1). When every 2nd slice was provided, the model demonstrated strong performance with DSC scores averaging 0.956 and 0.952 and HD values of 3.666 and 4.020 in the pre-operative and post-operative settings respectively (Table 2, Supplement eTable 1). However, as fewer ground truth slices were given (e.g., every 10th slice), DSC scores decreased to 0.856 in pre-operative cases and 0.751 in post-operative cases, while HD values increased (worse performance) to 7.450 and 15.36, respectively.

**Table 2.**
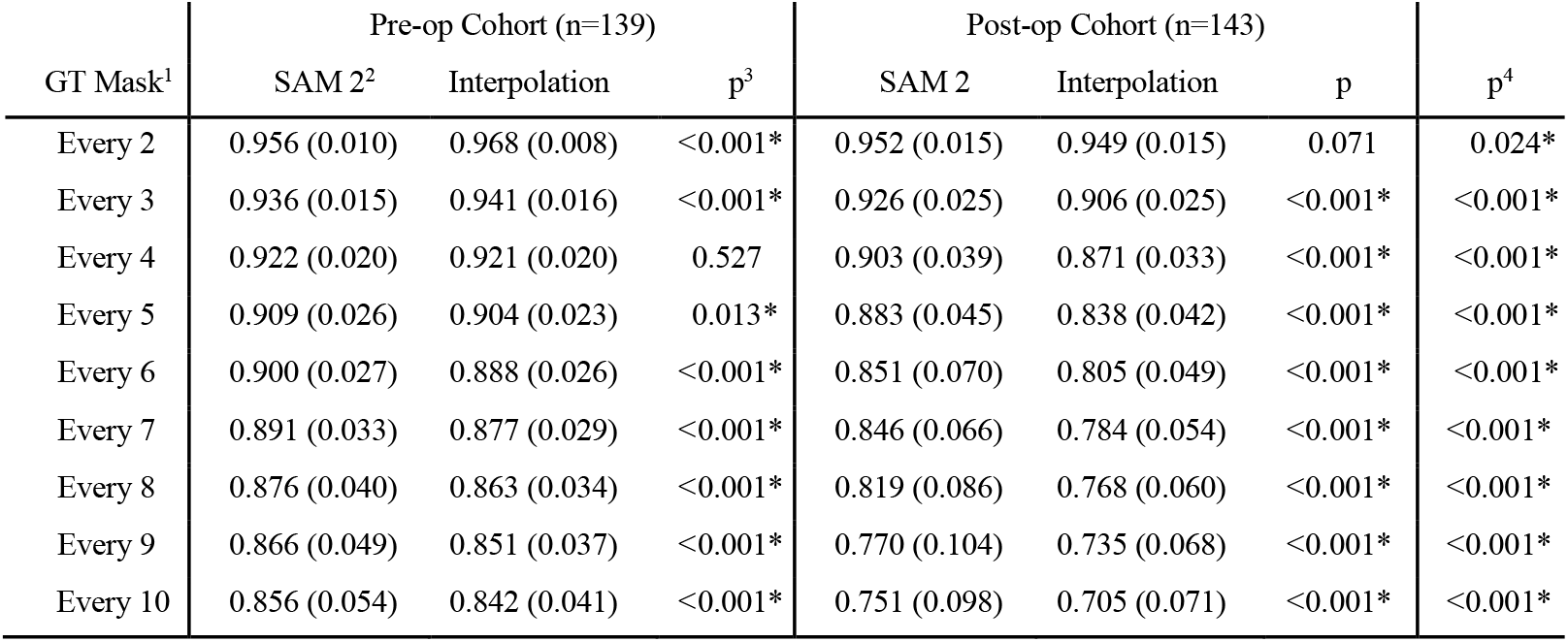
Dice similarity coefficient (DSC) for intact pre-operative and post-operative prostate cancer patient cases at varying levels of human segmentation.

**Figure 1.**
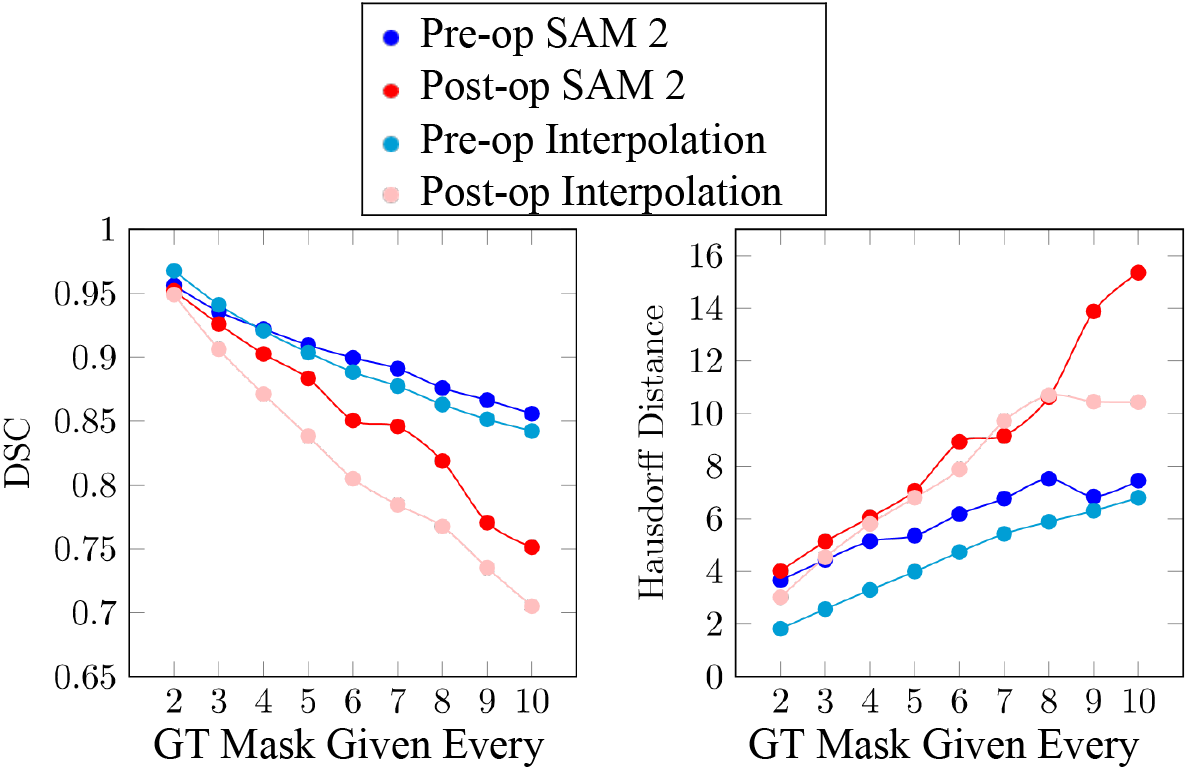
DSC scores and Hausdorff distances for foundation model performance for preoperative and post-operative prostate cancer radiotherapy target segmentation.

Overall, when fewer ground truth slices were provided, the SAM 2 model output and interpolation exhibited steadily decreasing performance on average for both DSC and HD. With decreased ground truth slices provided simulating reduced human intervention, SAM 2 outperformed interpolation and demonstrated better performance in the pre-operative setting compared to the post-operative setting in both DSC and HD metrics (Figure 1).

### DSC Performance

As the level of human intervention decreased (i.e. fewer ground truth slices provided), the average DSC for the SAM 2 model output in the pre-operative setting decreased from 0.956 to 0.856 for intervals ranging from every 2nd slice to every 10th slice (Table 2). Similarly, the DSC for interpolation decreased from 0.968 to 0.842 over the same interval. This trend highlights a consistent decline in AI segmentation accuracy as the spacing between ground truth slices widened. Notably, the pre-operative cases maintained higher DSC values than interpolation, with the performance gap widening at larger intervals. For example, at an interval of every 9th slice, the model achieved an average DSC of 0.866 (STD: 0.049), outperforming interpolation, which scored 0.851 (p < 0.001*).

In the post-operative cohort, the SAM 2 model also demonstrated a decline in DSC as the interval between ground truth slices increased. The average DSC decreased from 0.952 to 0.751 for intervals ranging from every 2nd slice to every 10th slice (Table 2). Interpolation in the post-operative setting showed a similar trend, with DSC decreasing from 0.949 to 0.705 over the same interval. The model consistently outperformed interpolation, also with the difference in performance widening at larger intervals. For example, at an interval of every 7th slice, the model achieved an average DSC of 0.846 (STD: 0.066), significantly outperforming interpolation, which scored 0.784 (p < 0.001*).

The pre-operative cases consistently achieved higher DSC values than the post-operative cases, particularly at lower levels of human intervention (i.e., larger intervals between ground truth slices). While both cohorts showed an approximately linear decrease in DSC as the interval increased, the decline was more pronounced in the post-operative setting (Table 2, Figure 2). For example, at an interval of every 10th slice, the pre-operative DSC of 0.856 was significantly higher than the post-operative DSC of 0.751 (p < 0.001*). Additionally, the standard deviation in DSC scores had a greater increase in the post-operative cohort (from 0.015 to 0.098) compared to the pre-operative cohort (from 0.010 to 0.054). These trends suggest that the SAM 2 model is more robust in the pre-operative setting, particularly when human intervention is minimal.

**Figure 2.**
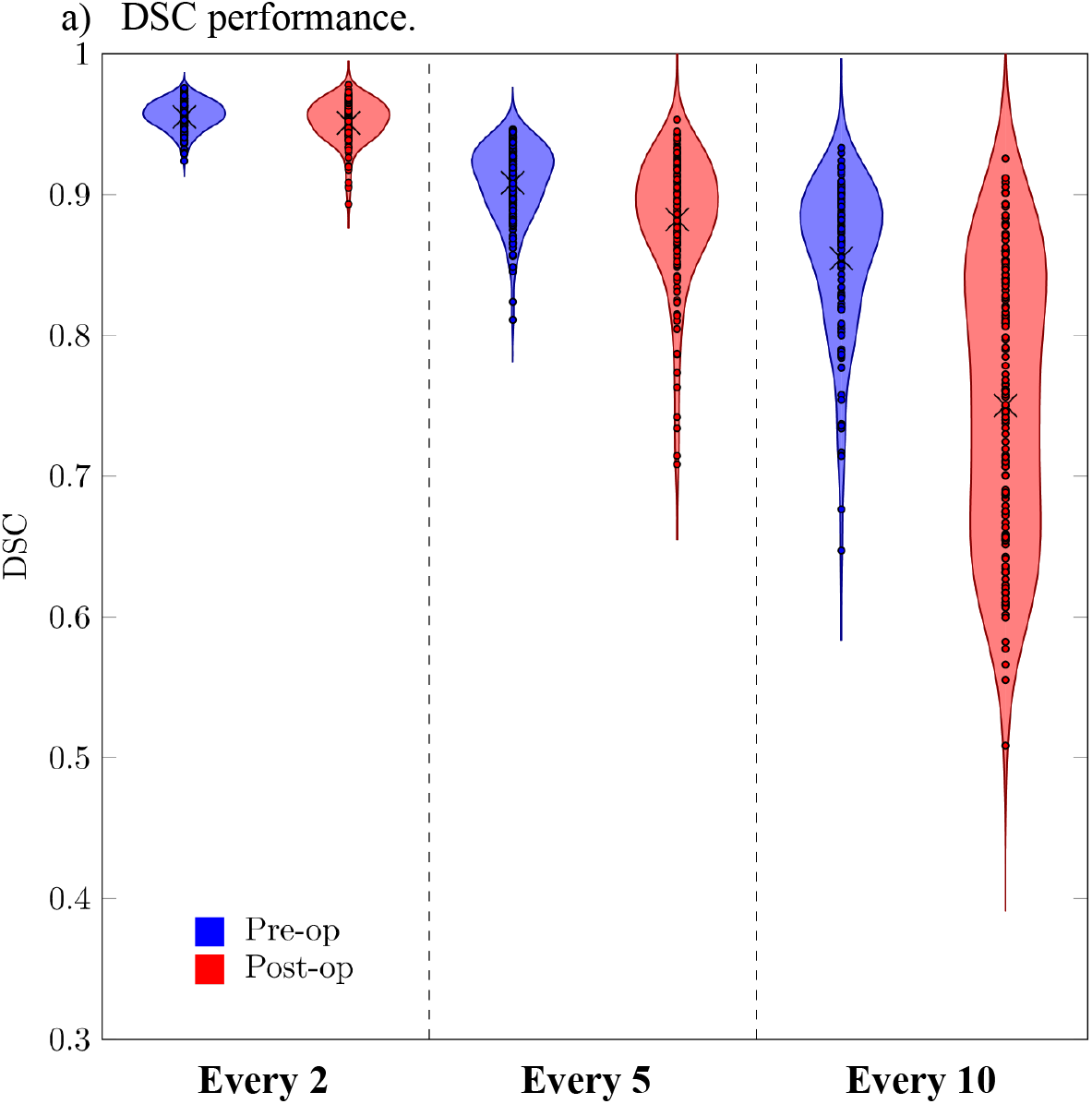
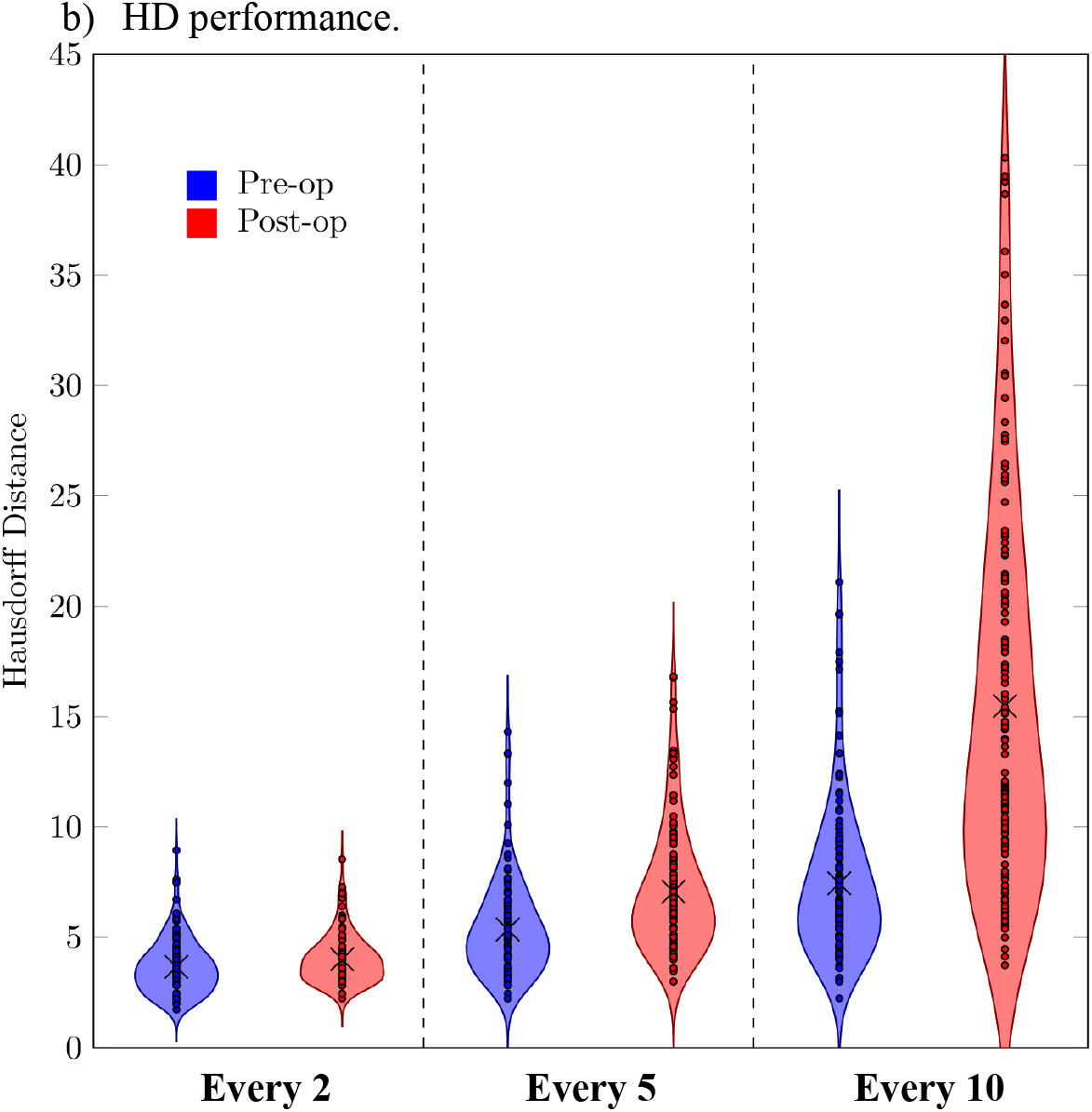
Violin plots of pre-operative and post-operative foundation model prostate cancer radiotherapy segmentation performance at selected intervals.

### HD Performance

The HD performance mirrored the DSC trends, with segmentation accuracy declining as the interval between ground truth slices increased. In the pre-operative setting, the average HD for the SAM 2 model increased from 3.666 to 7.450 (worse performance) for intervals ranging from every 2nd to every 10th slice. Similarly, in the post-operative cohort, the model’s HD increased from 4.020 to 15.36.

The pre-operative cases maintained lower HD values than the post-operative cases, with the performance gap widening at larger intervals. For example, at every 10th slice, the pre-operative HD of 7.450 was significantly lower than the post-operative HD of 15.36 (p < 0.001*). Variability also increased more sharply in the post-operative cohort, with standard deviations rising from 1.098 to 8.422 at intervals between every 2nd slice and every 10th slice, compared to 1.228 to 3.521 in the pre-operative cohort.

These findings corroborate the DSC results, demonstrating that segmentation accuracy and consistency deteriorate more significantly in the post-operative setting as human intervention decreases.

### Anatomical Position Analysis

For pre-operative cases, the superior part of the target segmentation had average DSC values ranging from 0.77 to 0.81 for the every 10th slice interval, 0.85 to 0.90 for every 5th slice, and 0.90 to 0.93 for every 2nd slice. The middle and inferior regions showed slightly higher segmentation accuracy at the every 10th slice interval, with DSC values ranging from 0.82 to 0.87, while the ranges of 0.85 to 0.91 for every 5th slice and 0.88 to 0.94 for every 2nd slice were in line with the superior region ranges (Figure 3). The difference in performance at the every 10th slice interval is likely reflective of seminal vesicles being present at the superior part of the intact CTV, leading to variations in ground truth and model outputs.

**Figure 3.**
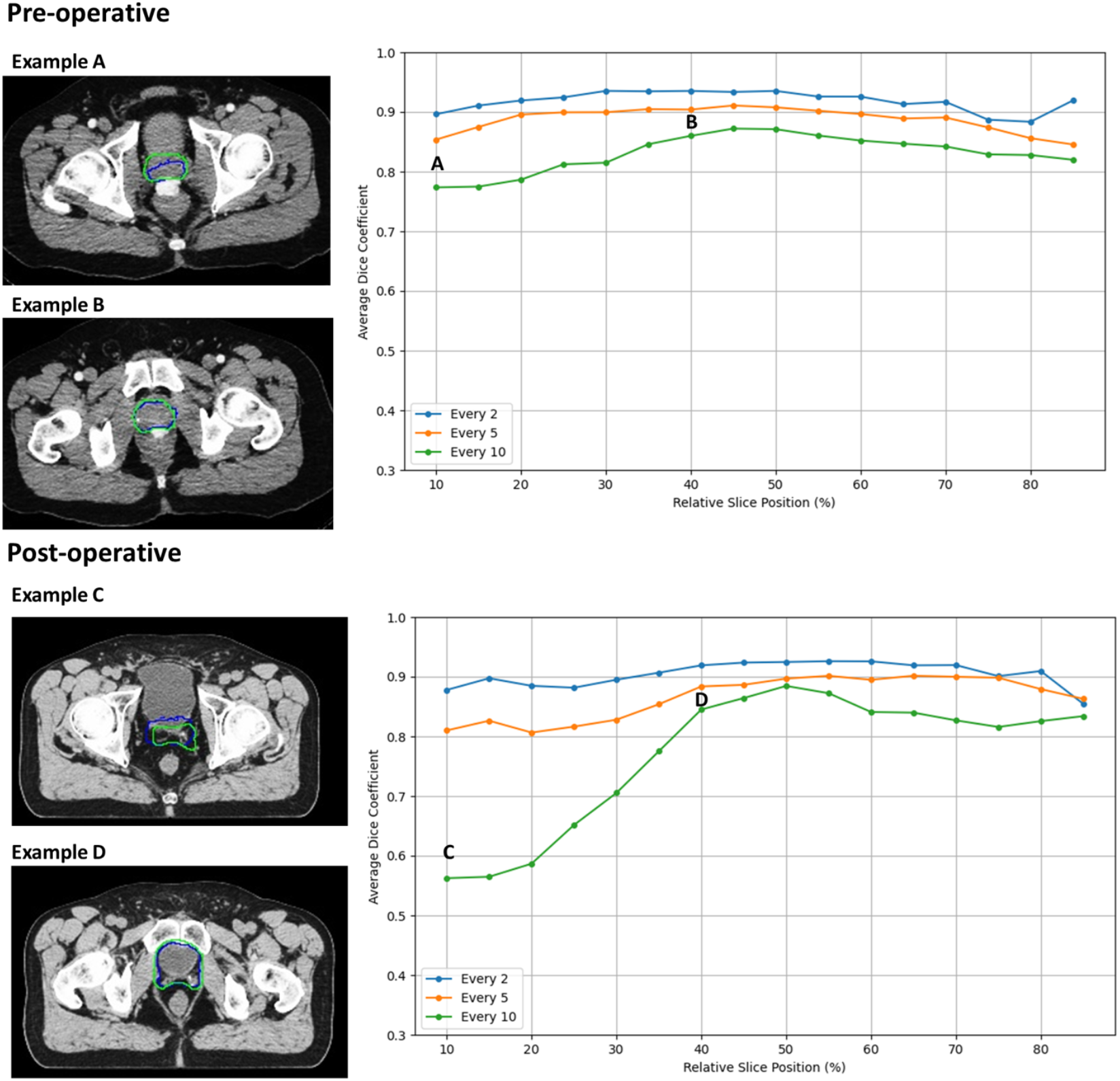
Foundation model pre-operative and post-operative prostate cancer radiotherapy segmentation DSC performance by relative anatomic position.

For post-operative cases, superior regions show average DSC values ranging from 0.56 to 0.71 for the every 10th slice interval, 0.81 to 0.83 for every 5th slice, and 0.87 to 0.90 for every 2nd slice (Figure 3). The middle and inferior regions achieve significantly better segmentation accuracy at the every 10th slice interval, with average DSC values ranging from 0.82 to 0.88, while the every 5th slice and every 2nd slice intervals ranged from 0.86 to 0.90 and 0.85 to 0.93 respectively. The more pronounced drop in target segmentation performance in the superior portion of the CTV within the post-operative cohort at the every 10th slice interval reflects a greater degree of variability and less well-defined boundaries than the pre-operative cases.

Figure 3 shows representative image pairs illustrating the anatomical dependence of segmentation accuracy. In superior regions of the CTV, AI predictions (blue) frequently deviated from the ground truth (green), particularly in areas involving complex anatomical structures such as the seminal vesicles or post-surgical changes in the prostate fossa. By contrast, middle and inferior regions exhibit closer alignment between predicted and ground truth contours, reflecting the model’s improved performance in areas with more distinct and stable anatomical boundaries.

## Discussion

Our study demonstrates that foundation models can achieve promising performance in prostate radiotherapy segmentation tasks with reduced human intervention and no domain-specific training. Unlike commercially available auto-segmentation tools designed specifically for this domain, SAM 2 offers a versatile, open-source solution that can be readily adapted to various medical imaging tasks. By testing the model on pre-operative and post-operative patient cases across a range of levels of human guidance, we provide a nuanced understanding of its capabilities across distinct clinical contexts. While SAM 2 does not outperform existing commercial prostate radiotherapy segmentation solutions, it maintained reasonable accuracy across varying levels of human guidance in pre-operative cases with clear anatomical boundaries.^26^

Our results highlight a promising paradigm shift in medical image analysis. Rather than developing specialized models for each clinical task, foundation models offer a more scalable approach through fine-tuning.^27,28^ This could significantly reduce development costs and accelerate the deployment of AI solutions across different medical imaging applications.^29^ This corroborates previous work with SAM 2 demonstrating feasibility in medical imaging, achieving superior performance through targeted modifications to the base model. While SAM 2 scored an average DSC of 0.86 on various anatomical structure segmentation tasks, a model specialized for medical imaging achieved an average of 0.89 through the use of a “self-sorting memory bank” that gives higher weight to more informative embeddings of the surrounding slices rather than giving the highest weight to the embeddings of the closest slices.^19^ These advancements demonstrate domain-specific adaptations can potentially improve foundation model performance without complete architectural redesign.

Despite these promising results, there are limitations to this approach. As a foundation model trained on general video and image data, SAM 2 is likely optimized to recognize clearly defined boundaries in everyday settings. However, in medical image analysis and radiotherapy segmentation, well-defined boundaries may not be representative of the data used to train the model and are not always present. A previous study found that SAM struggled to segment organs of interest with less well-defined boundaries mirroring our finding of worse performance on post-operative prostate fossa segmentation than intact prostate segmentation, particularly at lower levels of human intervention.^30^ These findings suggest that if foundation models like SAM 2 were to be adopted in medical image analysis and radiation segmentation, special attention should be given to scenarios that do not closely align with the data used to train the model.

There are other limitations to our study. This is a single institution analysis, with patients treated across a multiple year time period with shifts in practice patterns and radiotherapy technology. Our data was retrospectively collected with inherent limitations of such analysis. Nevertheless, the heterogeneity of the data may reflect potential of the foundation model in real-world settings. Future research will be needed to develop efficient fine-tuning strategies that can quickly adapt such foundation models to specific clinical tasks while preserving their generalization capabilities.^31–33^ This includes creating methods to incorporate domain expertise without requiring extensive retraining of the base model. Additionally, establishing robust frameworks for evaluating clinical reliability and safety will be crucial for healthcare adoption.^34,35^ If future models are found to be safe and reliable, integration with existing clinical workflows presents another key challenge, requiring careful consideration of user interface design, computational requirements, and compatibility with current clinical software systems.^36^ These developments will need to address both technical performance and practical implementation challenges to realize the full potential of foundation models in medical applications.

## Conclusions

Foundation models represent a significant AI advancement with promising applications for specialized medical segmentation tasks. For prostate cancer radiotherapy planning, its strengths were more evident in the pre-operative intact setting, where clear radiographic boundaries facilitate accurate segmentation with reduced human intervention. Further research is needed to refine such models for radiotherapy task-specific applications such as addressing challenges of post-operative segmentation. These advancements could pave the way for broader adoption of foundation models in medical imaging and personalized treatment planning.

## Supporting information

Supplement eTable 1

## Data Availability

All data produced in the present study are available upon reasonable request to the authors

## Footnotes

1 Physician-created ground truth mask provided as input every nth slice

2 DSC reported as mean (std)

3 P-value calculated by Mann-Whitney U test comparing SAM 2 and Interpolation performance within corresponding cohort

4 P-value calculated by Mann-Whitney U test comparing SAM 2 performance across cohorts

