## Supplement eTable 1 for "Performance of an artificial intelligence foundation model for prostate radiotherapy segmentation"

**Supplemental Table 1.** Hausdorff Distances for intact pre-operative and post-operative prostate cancer patient cases at varying levels of human segmentation.

| | Pre-op Cohort (n=139) | | | Post-op Cohort (n=143) | | | |
|---|---|---|---|---|---|---|---|
| GT Mask[1] | SAM 2[2] | Interpolation | p[3] | SAM 2 | Interpolation | p | p[4] |
| Every 2 | 3.666 (1.228) | 1.823 (0.843) | <0.001* | 4.020 (1.098) | 3.022 (1.293) | <0.001* | 0.004* |
| Every 3 | 4.439 (1.482) | 2.570 (0.985) | <0.001* | 5.136 (1.669) | 4.541 (1.807) | <0.001* | <0.001* |
| Every 4 | 5.148 (2.470) | 3.296 (0.882) | <0.001* | 6.054 (2.336) | 5.806 (2.163) | 0.402 | <0.001* |
| Every 5 | 5.359 (2.153) | 3.996 (0.756) | <0.001* | 7.070 (2.904) | 6.804 (2.212) | 0.851 | <0.001* |
| Every 6 | 6.179 (3.840) | 4.738 (0.742) | <0.001* | 8.927 (4.758) | 7.882 (2.562) | 0.423 | <0.001* |
| Every 7 | 6.769 (4.307) | 5.429 (1.380) | 0.072 | 9.155 (4.610) | 9.721 (3.153) | 0.005* | <0.001* |
| Every 8 | 7.521 (4.880) | 5.890 (1.170) | 0.128 | 10.63 (6.581) | 10.70 (3.072) | <0.001* | <0.001* |
| Every 9 | 6.845 (3.226) | 6.309 (1.225) | 0.857 | 13.89 (8.517) | 10.45 (2.961) | 0.057 | <0.001* |
| Every 10 | 7.450 (3.521) | 6.801 (1.534) | 0.980 | 15.36 (8.422) | 10.44 (2.503) | <0.001* | <0.001* |

[1] Physician-created ground truth (GT) mask provided as input every nth slice
[2] Hausdorff Distance reported as mean (std)
[3] P-value calculated by Mann-Whitney U test comparing SAM 2 and Interpolation performance within corresponding cohort
[4] P-value calculated by Mann-Whitney U test comparing SAM 2 performance across cohorts

### Example Case Comparison

Supplemental Figure 1 presents a visual comparison between SAM 2 predictions (blue contours) and ground truth (green contours) for representative pre-operative and post-operative cases across different slice intervals. Each row shows the segmentation at different anatomical levels (Up, Middle, and Bottom), allowing for detailed examination of model performance throughout the volume.

**Supplemental Figure 1.** Example case comparison of pre-operative and post-operative prostate target segmentation across various anatomical positions.

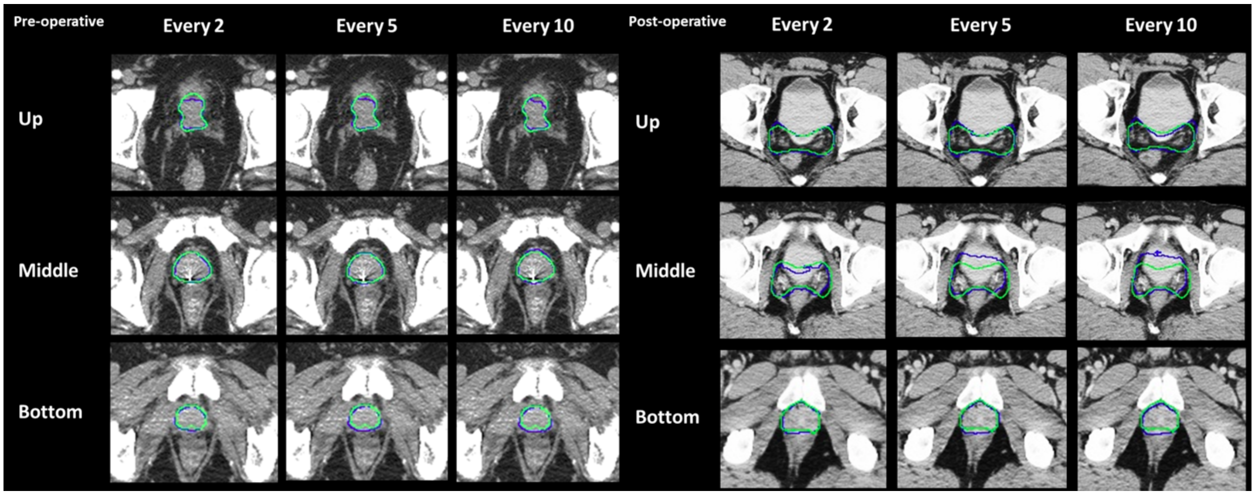


In the pre-operative case (left panel), SAM 2 demonstrates strong agreement with ground truth contours across all intervals, particularly in the middle slices where the prostate boundaries are most distinct. The model maintains consistent contour prediction even as the interval between ground truth slices increases from 2 to 10, though subtle deviations become apparent at the larger intervals, especially in the superior (Up) and inferior (Bottom) regions.

The post-operative case (right panel) reveals greater challenges in accurate segmentation. While SAM 2 captures the general shape of the prostate fossa, there are notable differences between predicted and ground truth contours, particularly in the middle portion where the surgical bed interfaces with the bladder. The superior and inferior slices show better agreement, though with more pronounced deviations compared to the pre-operative case. These visual findings align with the quantitative metrics, supporting the observation that SAM 2 performs more robustly in pre-operative cases where anatomical boundaries are better defined.

Of particular interest is the model's handling of complex anatomical interfaces in the post-operative setting. The middle slices demonstrate how SAM 2 sometimes struggles to precisely define the boundary between the prostate fossa and adjacent structures, especially at larger intervals. This visual evidence helps explain the higher HD values observed in post-operative cases, while still maintaining reasonable DSC scores due to overall volume agreement.
